# Summaries, Analysis and Simulations of Recent COVID-19 Epidemics in Mainland China

**DOI:** 10.1101/2022.04.03.22273225

**Authors:** Lequan Min

**Affiliations:** School of Mathematics and Physics, University of Science and Technology Beijing, Beijing 100083, PR China

**Keywords:** China COVID-19, infection transmission rates, infection blocking rates, recovery rates, modeling, simulations

## Abstract

**Background:** Globally COVID-19 epidemics have caused tremendous disasters. China prevented effectively the spread of COVID-19 epidemics before 2022. Recently Omicron and Delta variants cause a surge in reported COVID-19 infections.

**Methods:** Using differential equations and real word data, this study modelings and simulates COVID-19 epidemic in mainland China, estimates transmission rates, recovery rates, and blocking rates to symptomatic and asymptomatic infections. The transmission rates and recovery rates of the foreign input COVID-19 infected individuals in mainland China have also been studied.

**Results:** The simulation results were in good agreement with the real word data. The recovery rates of the foreign input symptomatic and asymptomatic infected individuals are much higher than those of the mainland COVID-19 infected individuals. The blocking rates to symptomatic and asymptomatic mainland infections are lower than those of the previous epidemics in mainland China. The blocking rate implemented between March 24-31, 2022 may not prevent the rapid spreads of COVID-19 epidemics in mainland China. For the foreign input COVID-19 epidemics, the numbers of the current symptomatic individuals and the asymptomatic individuals charged in medical observations have decreased significantly after March 17 2022.

**Conclusions:** Need to implement more strict prevention and control strategies to prevent the spread of the COVID-19 epidemics in mainland China. Keeping the present prevention and therapy measures to foreign input COVID-19 infections can rapidly reduce the number of foreign input infected individuals to a very low level.

## 1 Introduction

The 2019 coronavirus disease (COVID-19) has placed tremendous pressures onto the prevention, control, and healthcare systems worldwide. Many countries have experienced multiple outbreaks of the COVID-19 epidemic due to incomplete preventive measures and Omicron and Delta variants. As of 30 March 2022, there are more than 483.5 million confirmed cases of COVID-19 with more than 6.1 million deaths globally [1].

Modelling the dynamics of spread of disease can help people to understand the mechanism of epidemic diseases, formulate and evaluate prevention and control strategies, and predict tools for the spread or disappearance of an epidemic [2]. Since the outbreak of COVID-19 in Wuhan China, a large numbers of articles on modelings and predictions of COVID-19 epidemics have been published (for examples see [3–12]).

China prevent effectively the spread of COVID-19 epidemics before Omicron and Delta variants appeared. Recently, reported numbers of symptomatic and asymptomatic COVID-19 infected individuals are increased rapidly. This paper summarizes, analyzes and simulates the recent COVID-19 epidemic in mainland China, estimates infection transmission rates, infection blocking rates, and preventive measures through modelings and numerical simulations. The rest of this paper is organized as follows. Section 2 introduces materials and methods. Subsection 3.1 modelings and simulates the dynamics of COVID-19 Epidemics in Mainland. subsection 3.2 modelings and simulates the dynamics of foreign input COVID-19 Epidemics in China. Subsection 3.3 discuses and compares the simulation results. Virtual simulations for China Mainland COVID-19 epidemic and foreign input COVID-19 epidemic are implemented in the Subsections 3.4 and 3.5, respectively. Concluding remarks are given in Section 4.

## 2 Materials and Methods

The dataset of the China COVID-19 epidemics from December 31, 2021 to April 25, 2022 was collected and edited from the National Health Commission of the People’s Republic of China official website [13]. Using differential equation models stimulates the outcomes of the numbers of the current symptomatic individuals, the current asymptomatic individuals, the cumulative recovered symptomatic individuals and cumulative asymptomatic individuals discharged from observations. Equation parameters were determined by so-called minimization error square criterion described in references [12, 14, 15]. Using virtual simulations estimates outcomes of the spreads of COVID-19 epidemics in mainland China. Simulations and figure drawings were implemented via Matlab programs.

## 3 Analysis and Simulations of COVID-19 Epidemics in China

### 3.1 COVID-19 Epidemics in Mainland

Figure 1 shows the outcomes of the numbers of the current symptomatic individuals (CSI) and the current asymptomatic individuals (CAI). Figure 2 shows the outcomes of the numbers of the cumulative recovered symptomatic individuals (CCSI) and the cumulative asymptomatic individuals (CCAI) discharged from observations.

**Figure 1.**
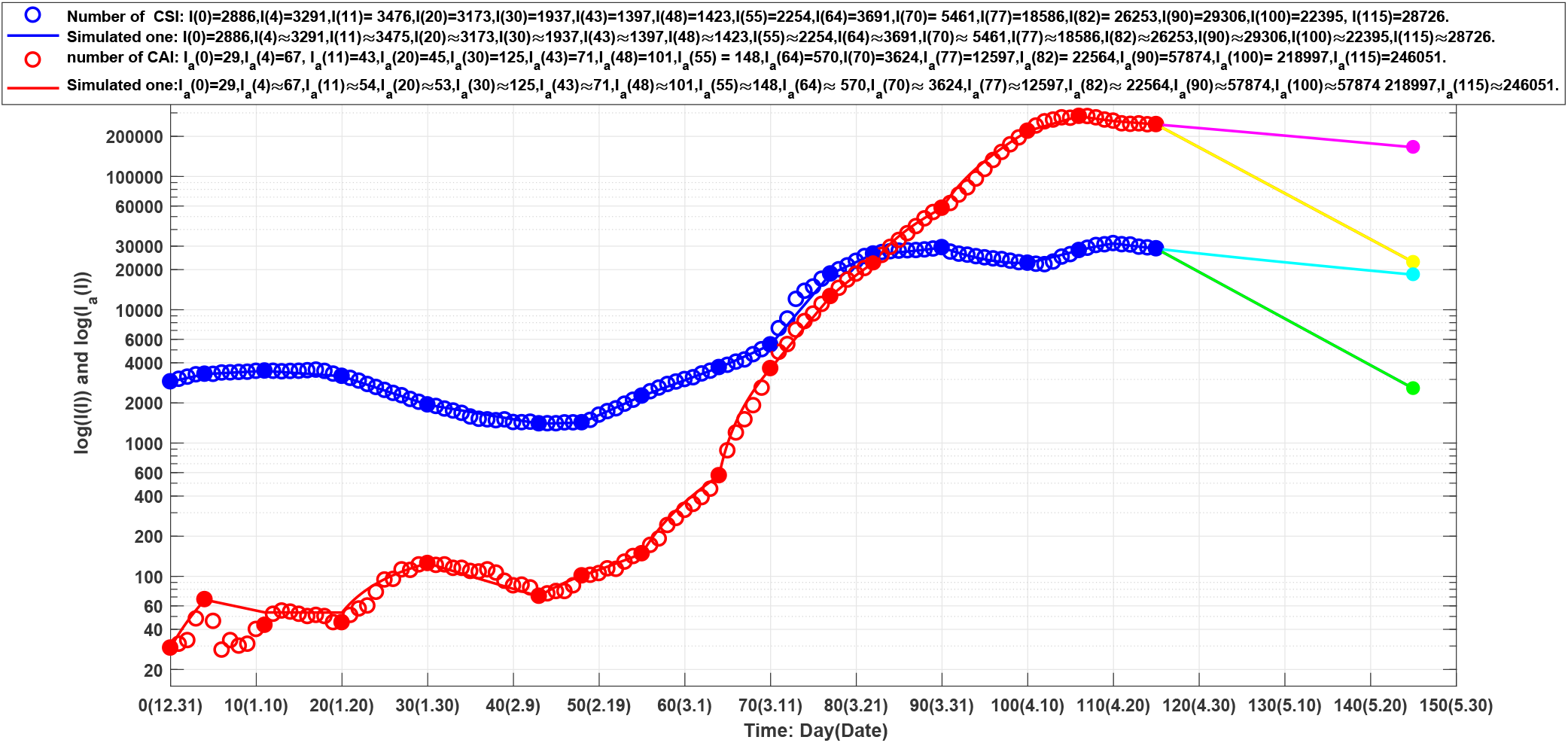
Blue circles: outcome of the number of the current symptomatic individuals (CSI), blue line: outcome of the corresponding simulations of equation (1). Red circles: outcome of the numbers of the current asymptomatic individuals (CAI) charged in medical observations, red line: outcome of the corresponding simulations of equation (1). The lines colored by cyan, magenta, green and yellow correspond to the virtual simulation results of equation (1). See Section Mainland Epidemic Virtual Simulations for details.

**Figure 2:**
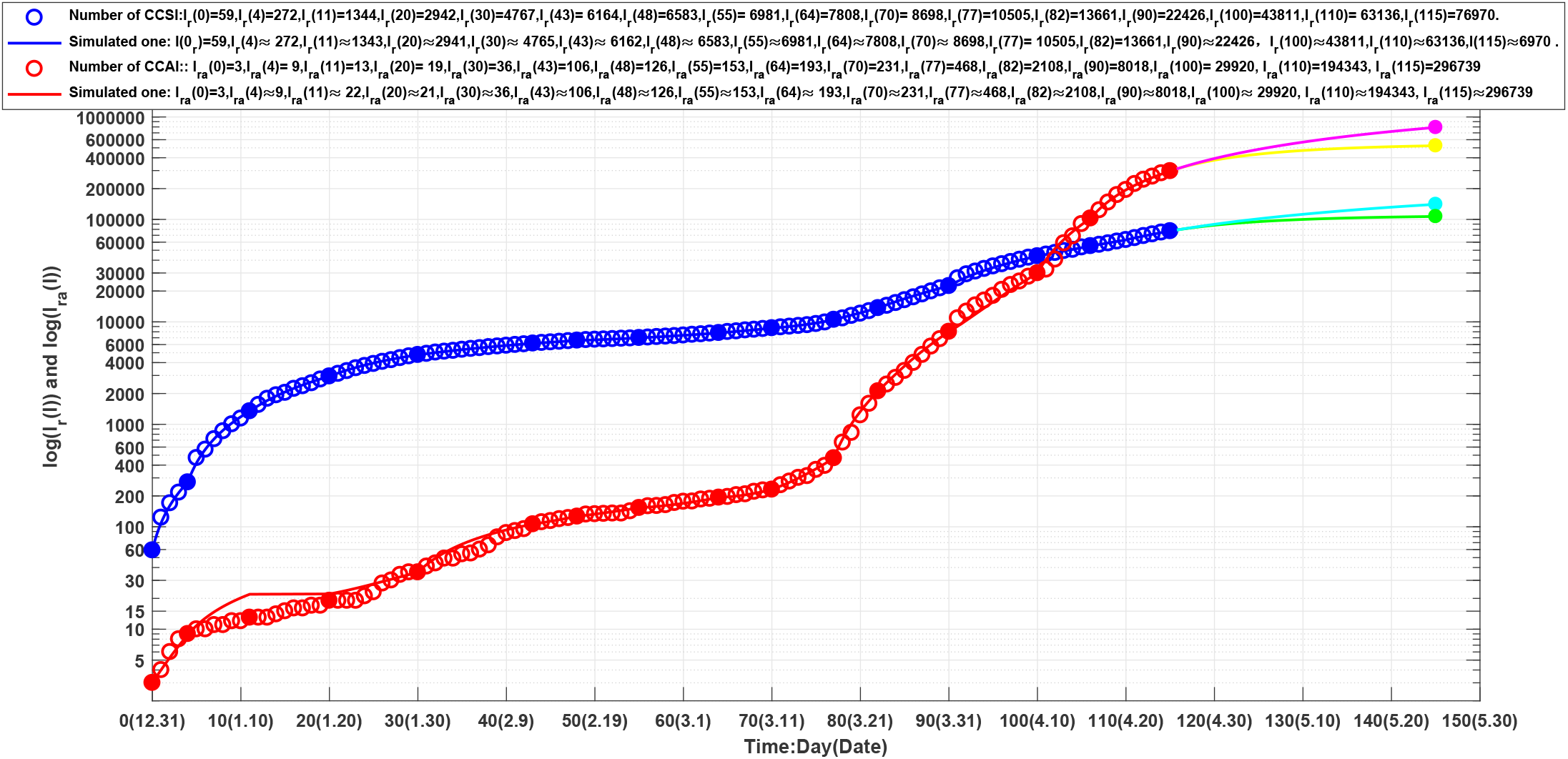
Blue circles: outcome of the number of the current cumulative recovered symptomatic individuals (CCSI), blue line: outcome of the corresponding simulations of equation (1). Red circles: outcome of the numbers of the current cumulative asymptomatic individuals (CCAI) discharged in medical observations, red line: outcome of the corresponding simulations of equation (1). The lines colored by cyan, magenta, green and yellow correspond to the virtual simulation results of equation (1). See Section Mainland Epidemic Virtual Simulations for details.

The recent COVID-19 epidemics in mainland China are still continuing. Although there existed a turning point of the current symptomatic infections on day 17 (January 17, 2021), after day 43 (February 12, 2022), numbers of the the current symptomatic and asymptomatic infections are increasing until reached to the second turning point on day 110 (April 20). For the current asymptomatic infections charged i medical observations, the number were increasing until reached recently to the first turning point on day 106 (April 16).

In order to estimate numerically the transmission rates and blocking rates to symptomatic and asymptomatic infections, we need to set up mathematic models (similar to [12, 14, 15]) to simulate the dynamics of spread of infection disease. Because we modeling uniformly the epidemic situations appeared in different provinces and regions, the transmission rates *β*_*ij*_ of symptomatic infections and asymptomatic infections are changing over different transmission intervals.

For the mainland epidemics over the *lth* transmission interval, the symptomatic infected individuals (*I*) and the asymptomatic infected individuals (*I*_*a*_) infect the susceptible population (*S*) with the transmission rates of *β*_11_(*l*) and *β*_21_(*l*), respectively, making *S* become symptomatic infected individuals, and with the transmission rates of *β*_12_(*l*) and *β*_22_(*l*), respectively, making *S* become asymptomatic individuals. Then, a symptomatic individual is cured at a rate *κ*(*l*), an asymptomatic individual returns to normal at a rate *κ*_*a*_(*l*). Here all parameters are positive numbers. Assume that the dynamics of an epidemic can be described by *m*-time intervals, which correspond different transmission rates, prevention and control measures, and medical effects. At *l*th time interval, the model has the form (similar to [12, 14, 15]):

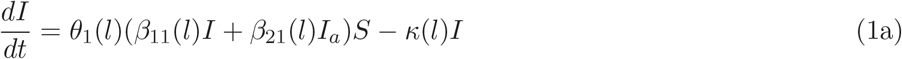

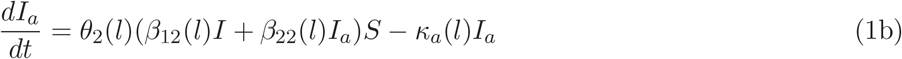

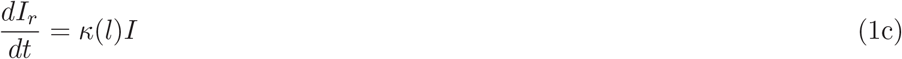

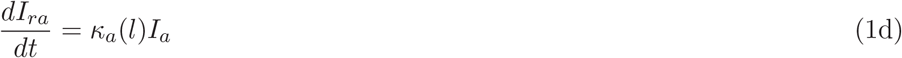

where Θ_1_(*i*) = (1 − *θ*_1_(*l*)) and Θ_2_(*l*) = (1 − *θ*_2_(*l*)) (*l* = 1, …, *m*) represent the blocking rates to symptomatic and asymptomatic infections, respectively.

It can be assumed that the input transmissions can be divided into 16 time intervals (see solid points in Figs. 1 and 2). We need to determine the parameters of equation (1) for *l* = 1, 2, …, 16. Over the *lth* time interval [*t*_*l*−1_, *t*_*l*_],

Denote

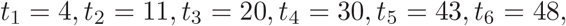

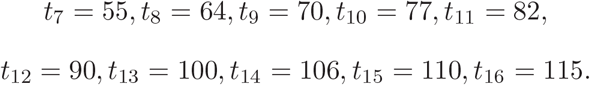

Denote *I*_*c*_(*t*_*l*_) to be the number of the reported current symptomatic infected individuals, and *I*_*ca*_(*t*_*l*_) be the number of the reported current asymptomatic individuals charged in medical observations. Denote 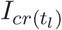 to be the number of the reported current cumulative recovered symptomatic infected individuals, and *I*_*cra*_(*t*_*l*_) be the number of the reported current cumulative asymptomatic individuals discharged in medical observations. Using the minimization error square criterion:

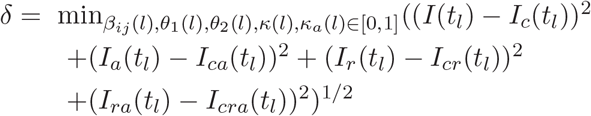

determines the *β*_*ij*_(*l*)^′^*s, κ*(*l*)^′^*s, κ*_*a*_(*l*)^′^*s*), *θ*_1_(*l*)^′^*s*, and *θ*_2_(*l*)^′^*s*. The calculated parameters are shown in Table 1. The corresponding simulation results of equation (1) are shown in Fig. 1 and Fig. 2. Observe that the simulation results of equation (1) were in good agreement with the data of the COVID-19 epidemics in mainland China (see the solid blue lines, the red lines in Fig. 1 and Fig. 2).

**Table 1.** Equation parameters of the COVID epidemics in mainland China during 2021.12.31-2022.4.25.

| $l$ | Days | $\beta_{11}(l)$ | $\beta_{21}(l)$ | $\beta_{12}(l)$ | $\beta_{22}(l)$ | $\Theta_1(l)$ | $\Theta_2(l)$ | $\kappa(l)$ | $\kappa_a(l)$ |
| --- | --- | --- | --- | --- | --- | --- | --- | --- | --- |
| 1 | 0-4 | 0.049056 | 0.072052 | 0.002102 | 0.094068 | 0% | 0% | 0.017296 | 0.03107000 |
| 2 | 5-11 | 0.058000 | 0.072052 | 0.000100 | 0.000100 | 10.6% | 99% | 0.045215 | 0.03107017 |
| 3 | 12-20 | 0.056600 | 0.072052 | 0.002100 | 0.000094 | 25% | 99% | 0.053450 | 0.00031700 |
| 4 | 21-30 | 0.044400 | 0.072052 | 0.004100 | 0.003990 | 49.84% | 19.5% | 0.073030 | 0.01485000 |
| 5 | 31-43 | 0.077340 | 0.071000 | 0.002500 | 0.003 | 51.02% | 72.5% | 0.065000 | 0.05670000 |
| 6 | 44-48 | 0.124800 | 0.071000 | 0.025000 | 0.003 | 51.15% | 71.6% | 0.059440 | 0.04670000 |
| 7 | 49-55 | 0.124800 | 0.071000 | 0.020900 | 0.003 | 25.02% | 72.3% | 0.031480 | 0.03140000 |
| 8 | 56-64 | 0.124900 | 0.017800 | 0.059000 | 0.003 | 31.98% | 70.29 % | 0.031550 | 0.01290000 |
| 9 | 65-70 | 0.125200 | 0.017800 | 0.141000 | 0.009 | 26.02% | 21.07% | 0.032940 | 0.00319000 |
| 10 | 71-77 | 0.287270 | 0.1 | 0.160630 | 0.009 | 43.846% | 26.314% | 0.024102 | 0.00468900 |
| 11 | 78-82 | 0.287270 | 0.1 | 0.160630 | 0.009 | 73.264% | 37.508% | 0.028488 | 0.01894100 |
| 12 | 83-90 | 0.287270 | 0.1 | 0.294230 | 0.009 | 87.601% | 39.036% | 0.039846 | 0.01839700 |
| 13 | 91-100 | 0.287200 | 0.1 | 0.5 | 0.1 | 92.89% | 28.441% | 0.087194 | 0.01645073 |
| 14 | 101-106 | 0.287200 | 0.1 | 0.5 | 0.1 | 91.337% | 39.282% | 0.074840 | 0.04838250 |
| 15 | 106-110 | 0.287270 | 0.1 | 0.5 | 0.1 | 91.846% | 58.691 | 0.068019 | 0.08472660 |
| 16 | 110-115 | 0.287270 | 0.1 | 0.5 | 0.1 | 93.439% | 56.636 | 0.0921248 | 0.08093270 |

### 3.2 Foreign input COVID-19 Epidemics in China

Figure 3 shows outcomes of the numbers of the current symptomatic individuals (CSI) and the current asymptomatic individuals (CAI). Figure 4 shows outcomes of the numbers of the cumulative recovered symptomatic individuals (CCSI) and the cumulative asymptomatic individuals (CCAI) discharged from observations.

**Figure 3:**
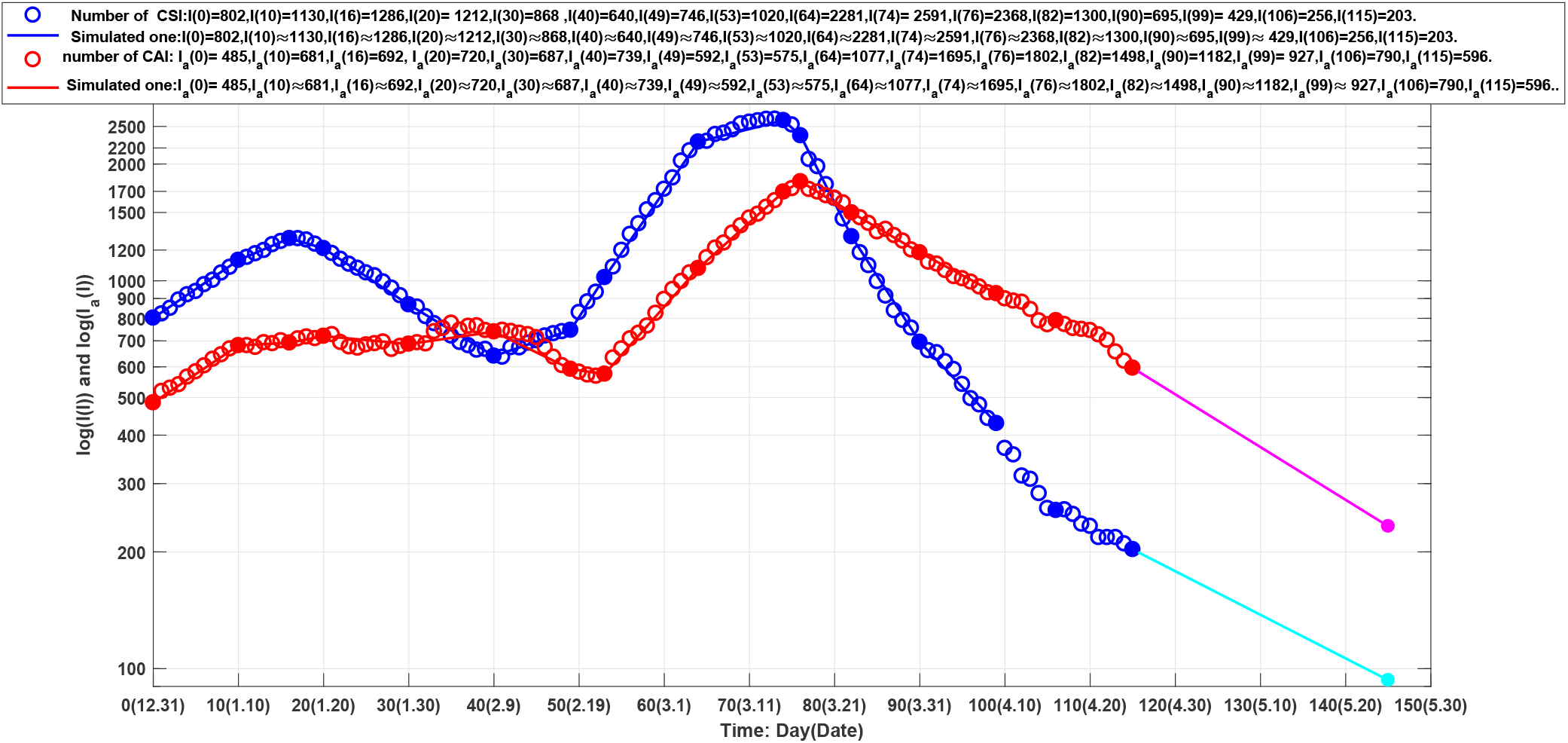
Blue circles: outcome of the number of the current symptomatic individuals (CSI), blue line: outcome of the corresponding simulations of equation (2). Red circles: outcome of the numbers of the current asymptomatic individuals (CAI) charged in medical observations, red line: outcome of the corresponding simulations of equation (2). The lines colored by cyan and magenta correspond to the virtual simulation results of equation (2). See Section Foreign Input Epidemic Virtual Simulations for details.

**Figure 4:**
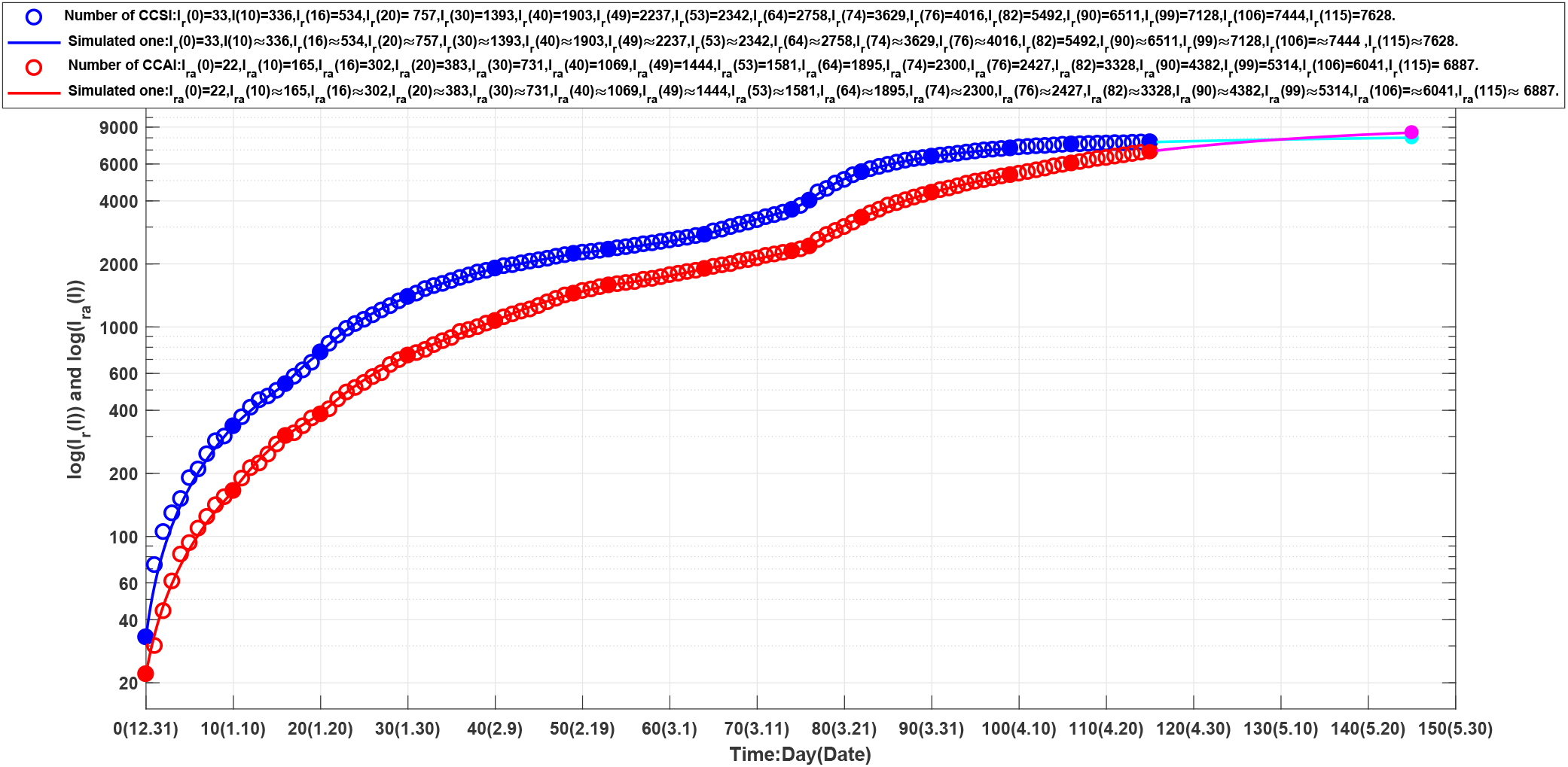
Blue circles: outcome of the number of the current cumulative recovered symptomatic individuals (CCSI), blue line: outcome of the corresponding simulations of equation (2). Red circles: outcome of the numbers of the current cumulative asymptomatic individuals (CCAI) discharged in medical observations, red line: outcome of the corresponding simulations of equation (2). The lines colored by cyan and magenta correspond to the virtual simulation results of equation (2). See Section Foreign Input Epidemic Virtual Simulations for details.

The epidemics have reached the first turning point of the current symptomatic infected individuals on day 16. After day 40, the number of the current symptomatic individuals began to increase until on day 74 (March 15) to reach the second infection turning point.

The number of the current asymptomatic individuals charged in medical observations reached the first infection turning point on day 35. After day 52, the number began to increase until on day 76 (March 17) to reach the second infection turning point.

For the foreign input COVID-19 infected individuals, they were discovered immediately and no further transmissions generated each other after entering China. Therefore the model has simply the form

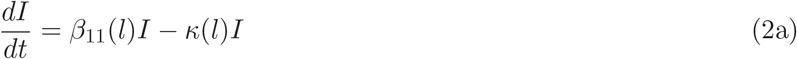

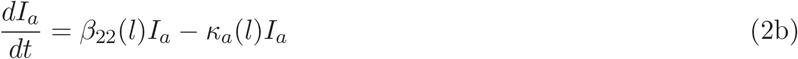

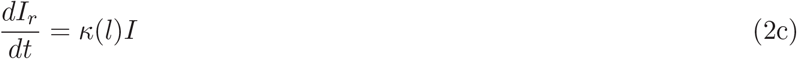

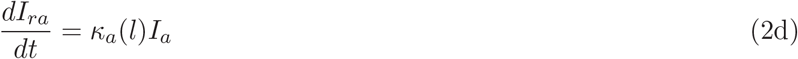

where *β*_11_(*l*) and *β*_22_(*l*) represent input transmission rates of the symptomatic individuals and asymptomatic individuals over the *lth* time interval, respectively.

It can be assumed that the input transmissions can be divided into 15 time intervals (see solid points in Fig. 3 and Fig. 4). We need to determine the parameters for equation (2) for *l* = 1, 2, …, 15 over the *lth* time interval [*t*_*l*−1_, *t*_*l*_].

Denote

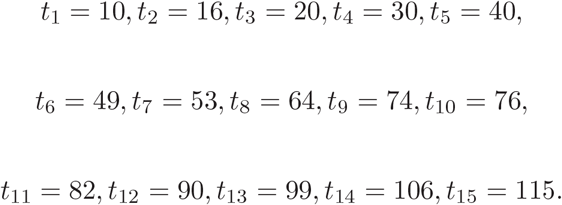

Denote *I*_*c*_(*t*_*l*_) to be the number of the reported current symptomatic infected individuals, and *I*_*ca*_(*t*_*l*_) be the number of the reported current asymptomatic individuals charged in medical observations. Denote 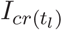 to be the number of the reported current cumulative recovered symptomatic infected individuals, and *I*_*cra*_(*t*_*l*_) be the number of the reported current cumulative asymptomatic individuals discharged from medical observations.

Using the minimization error square criterion:

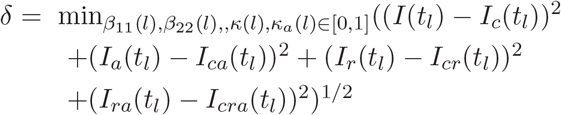

determines the *β*_11_(*l*)^′^*s, β*_22_(*l*)^′^*s, κ*(*l*)^′^*s* and *κ*_*a*_(*l*)^′^*s*. The calculated parameters are shown in Table 2. The corresponding simulation results of equation (2) are shown in Fig. 3 and Fig. 4. Observe that the simulation results of equation (2) were in good agreement with the data of the foreign input COVID-19 epidemics (see the solid blue lines and the red lines in Fig. 3 and Fig. 4). On the end points of the 15 investigated time-interval [*t*_*l*−1_, *t*_*l*_]^′^*s*, the simulated numbers and the actual reported numbers were approximate the same–errors were less than one person, respectively.

**Table 2.** Equation parameters of the foreign input COVID epidemics in mainland China during 2021.12.31-2022.4.25.

| $l$ | Days | $\beta_{11}(l)$ | $\beta_{22}(l)$ | $\kappa(l)$ | $\kappa_a(l)$ |
| --- | --- | --- | --- | --- | --- |
| 1 | 0-10 | 0.065980 | 0.058698 | 0.031686 | 0.024732 |
| 2 | 11-16 | 0.048874 | 0.036028 | 0.027328 | 0.033369 |
| 3 | 17-20 | 0.029875 | 0.038568 | 0.044693 | 0.028647 |
| 4 | 21-30 | 0.028319 | 0.044700 | 0.061721 | 0.049416 |
| 5 | 31-40 | 0.037650 | 0.054748 | 0.068147 | 0.047444 |
| 6 | 41-49 | 0.070788 | 0.038286 | 0.053722 | 0.062917 |
| 7 | 50-53 | 0.108306 | 0.051267 | 0.030017 | 0.058598 |
| 8 | 54-64 | 0.097254 | 0.092713 | 0.024111 | 0.035653 |
| 9 | 65-74 | 0.048546 | 0.075085 | 0.035803 | 0.029734 |
| 10 | 75-76 | 0.033113 | 0.066995 | 0.078092 | 0.036350 |
| 11 | 77-82 | 0.038166 | 0.060467 | 0.138110 | 0.091284 |
| 12 | 83-90 | 0.053600 | 0.069150 | 0.131850 | 0.098770 |
| 13 | 91-99 | 0.070690 | 0.071710 | 0.124300 | 0.098700 |
| 14 | 100-106 | 0.060960 | 0.098300 | 0.134700 | 0.121200 |
| 15 | 107-115 | 0.063440 | 0.105280 | 0.089306 | 0.136590 |

### 3.3 Results and Discussions

#### 3.3.1 Mainland Epidemics

Recent China COVID-19 epidemics with both Omicron and Delta variants make more difficult to prevent spread of the diseases. On the end points of the twelve investigated time-interval [*t*_*l*−1_, *t*_*l*_]^′^*s*, that is on day 0, day 4, day 11, day 20, day 30, day 43, day 48, day 55, day 64, day 70 day 77 day 82, day 90, day 100, day 106, day 110 and day 115, we obtain the following results (also see Figs. 1 and 2).

- On day 11, there was only 1 difference between the numbers of the reported and the simulated current symptomatic individuals. On the other end point days, the numbers of the reported and the simulated current symptomatic individuals were approximate the same.
- On day 11 and day 20, there were 11 and 8 differences between the numbers of the reported and the simulated current asymptomatic individuals charged in observations. On the other end point days, the numbers of the reported and the simulated current asymptomatic individuals charged in observations were approximate the same.
- On day 11, day 20, day 30, and day 43, there were only one or two differences between the numbers of the reported and the simulated current cumulative recovered symptomatic individuals. On the other end point days, the numbers of the reported and the simulated current cumulative recovered symptomatic individuals were approximate the same.
- On day 11 and day 20, there were nine and three differences between the numbers of the reported and the simulated current cumulative asymptomatic individuals discharged from observations. On the other end point days, the numbers of the reported and the simulated current cumulative asymptomatic individuals discharged from observations were approximate the same.
- The transmission rates of the symptomatic infections caused by the symptomatic individuals were increasing from day 21 (January 20) to day 70 (March 10), and then seems to stop (see *β*_11_(*l*)^′^*s* in Table 1).
- The transmission rates of the asymptomatic infections caused by the symptomatic individuals have obviously increased after day 56 (February 24, see *β*_12_(*l*)^′^*s* in Table 1).
- The transmission rates of the symptomatic infections caused by the asymptomatic individuals have obviously increased after day 55(see *β*_21_(*l*)^′^*s* in Table 1).
- The transmission rates of the symptomatic infections caused by the asymptomatic individuals have obviously increased after day 55 (see *β*_21_(*l*)^′^*s* in Table 1).
- During the first 70 days (before March 30), the transmission rates of the asymptomatic infections caused by the asymptomatic individuals are very low (see *β*_22_(*l*)^′^*s* in Table 1). After day 70, the transmission rates increased significantly.
- The blocking rates Θ_1_(*l*)^′^*s* and Θ_2_(*l*)^′^*s* to symptomatic and asymptomatic infections were not hight. Even on day 90 (March 30), the blocking rates only reach about 87.6% and 39.0% (see Table 1), respectively. However, for the first and second epidemics in Beijing and the five wave epidemics in Shanghai, the blocking rates reached to over 95% in one month [14–16].
- The recovery rates *κ*(*l*) and *κ*_*a*_(*l*) of the symptomatic individuals and asymptomatic individuals waved. The recovery rates reached first maximums during days 21-30 and days 31-43, respectively. In the last two weeks (After day 100, April 9), The recovery rates increased significantly (see Table 1).

#### 3.3.2 Foreign input epidemics

It seems that the foreign input COVID-19 infected individuals have been obtained good managements and therapies. On the end points of the 15 investigated time-interval [*t*_*l*−1_, *t*_*l*_]^′^*s*, that is on day 0, day 10, day 16, day 20, day 30, day 40, day 49, day 53, day 64, day 74 day 76 day 82 day 90, day 99, day 106, and day 115, we obtain the following results (also see Figs. 3 and 4).

1. On the end points of the 15 investigated time-interval [*t*_*l*−1_, *t*_*l*_]^′^*s*, the numbers of the reported and the simulated current symptomatic individuals were approximate the same.
2. On the end points of the 15 investigated time-interval [*t*_*l*−1_, *t*_*l*_]^′^*s*, the numbers of the reported and the simulated current asymptomatic individuals charged in medical observation were approximate the same.
3. On the end points of the 15 investigated time-interval [*t*_*l*−1_, *t*_*l*_]^′^*s*, the numbers of the reported and the simulated current cumulative recovered symptomatic individuals were approximate the same.
4. On the end points of the 15 investigated time-interval [*t*_*l*−1_, *t*_*l*_]^′^*s*, the numbers of the reported and the simulated current cumulative asymptomatic individuals discharged from medical observations were approximate the same.

- The input transmission rates of the symptomatic infection individuals and the asymptomatic infection individuals waved. The maximal input transmission rates reached during days 50-64, and days 54-64, 100-115 respectively (see Table 2).
- The recovery rates *κ*(*l*)^′^*s* of the symptomatic individuals were increasing during days 11-40 and days 54-82. The recovery rates *κ*_*a*_(*l*)^′^*s* of the asymptomatic individuals were increasing during days 17-49, and days 65-90 (see Table 2).
- After days 74 (March 14) and 76 (March 16, 2022), the turning point of the numbers of the current symptomatic individuals and the current asymptomatic individuals charged in observations appeared, respectively.

#### 3.3.3 Comparing

- The average recovery rate of the foreign input COVID-19 symptomatic infected individuals was about 0.071572. The average recovery rates of the mainland COVID-19 infected asymptomatic individuals was about 0.051501. The average recovery rate of the foreign input individuals was much higher than the one of the mainland people (see *κ*(*l*)^′^*s* in Table 1 and Table 2).
- The average recovery rate of the foreign input COVID-19 asymptomatic infected individuals was about 0.06356. The average recovery rate for mainland COVID-19 infected asymptomatic individuals was about 0.03131. The average recovery rate of the foreign input individuals was much higher than the one of the mainland people (see *κ*_*a*_(*l*)^′^*s* in Table 1 and Table 2).
- The average input transmission rate of the foreign input symptomatic infection individuals was about 0.057037, which was much lower than the average transmission rate 0.17474 of the symptomatic infection causing by the mainland symptomatic individuals (see *β*_11_(*l*)^′^*s* in Table 1 and Table 2).
- The average input transmission rate of the foreign input asymptomatic infection individuals was about 0.064133, which was much higher than the average transmission rate 0.034141. of the asymptomatic infection causing by the mainland symptomatic individuals (see *β*_12_(*l*)^′^*s* in Table 1 and *β*_22_(*l*)^′^*s* in Table 2).
- The last 24 days’ prevention measures implemented for mainland COVID-19 infected individuals reduced significantly the spread speed of the symptomatic infections (see Fig. 1 and Table 1). However it needs more effective prevention measures such that the numbers of the infected symptomatic and asymptomatic individuals decrease more rapidly.
- During days 83 to 115, the ratio of the average blocking rates of the asymptomatic infection and the symptomatic infection of the mainland epidemic was about 0.4958. Therefor it needs to increase largely the blocking rates to the asymptomatic infection.

## 4 Virtual Simulations

### 4.1 Mainland Epidemic Virtual Simulations

Assume that after day 115 (April 24, 2022), it still keeps the transmission rates 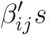 *s*, the blocking rates (Θ_1_(16), Θ_2_(16)), the recovery rates *κ*(16), and *κ*_*a*_(16) until day 145 (May 25, 2022). The simulation results of equation (1) are shown in Fig. 1 and Fig. 2 by cyan lines and magenta lines, respectively. Calculated results show that on day 145 (May 25), the numbers of current symptomatic and the asymptomatic infected individuals reach about 18329 and 1165535, respectively. The numbers of cumulative recovered symptomatic individuals and cumulative asymptomatic individuals discharged in medical observations reach about 140452 and 140452, respectively.

Furthermore assume that after day 115, it still keeps the transmission rates 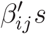, and recovery rates *κ*(16) and *κ*_*a*_(16) but increases the blocking rates to (Θ_1_, Θ_2_) ≡ (99%, 99%) until day 145. The simulation results of equation (1) are shown in Fig. 1 and Fig. 2 by green lines and yellow lines, respectively. Calculated results show that on day 145, the numbers of current symptomatic and the asymptomatic infected individuals reach about 2564 and 22708, respectively. The numbers of cumulative recovered symptomatic individuals and cumulative asymptomatic individuals discharged in medical observations reach about 106872 and 524005 respectively.

### 4.2 Foreign Input Epidemic Virtual Simulations

Assume that after day 115 (April 24, 2022), it still keeps the transmission rates *β*_15_(15), *β*_22_(11), the recovery rates *κ*(11), and *κ*_*a*_(11) until day 145 (25 May, 2022). The simulation results of equation (2) are shown in Fig. 3 and Fig. 4 by cyan lines and magenta lines, respectively. Calculated results show that on day 145, the numbers of the current symptomatic and the asymptomatic infected individuals reduce to 93 and 233, respectively. The numbers of cumulative recovered symptomatic individuals and cumulative asymptomatic individuals discharged from medical observations reach about 8006 and 8470, respectively.

## 5 Concluding Remarks

The main contributions of this paper are summarized as follows:

1. It is the first time to summary the COVID-19 epidemic from December 31 2021 to April 24, 2022 in mainland China. It shows a clear picture to prevent and control the spread of the COVID-19 China epidemics [13].
2. It uses two models to simulate the recent China epidemics. The simulation results on the end points of transmission intervals were in good agreement with the real word data [13], in particular the case for foreign input infections.
3. The simulation results can provide possible interpretations and estimations of the prevention and control measures, and the effectiveness of the treatments.
4. The recovery rates of the foreign input symptomatic and asymptomatic infected individuals were much higher than those of the mainland COVID-19 infected individuals.
5. Virtual simulations suggest that

- The evolution of the number of the current symptomatic individuals may be located in the region between the cyan line and the green line shown in Fig. 1 if the recovery rates and treatment efficacy keep the levels on April 24, 2022, depending on practical blocking rates to infections. Similarly, the evolutions of the number of the current asymptomatic individuals charged in medical observations may be located in the region between the magenta line and the yellow line shown in Fig. 1.
- The evolution of the number of the current cumulative recovered symptomatic individuals may be located in the region between the cyan line and the green line shown in Fig. 2. The evolutions of the number of the current cumulative asymptomatic individuals discharged in medical observations may be located in the region between the magenta line and the yellow line shown in Fig. 2.
- Although the infection turning points have appeared, it needs to rise the recovery rates *κ*(*l*)^′^*s, κ*_*a*_(*i*)^′^*s* and the blocking rates Θ_1_(*l*)^′^*s* and Θ_2_(*i*)^′^*s* of the mainland COVID-19 infected individuals to reduce the spreads of the COVID-19 epidemics.
- Keeping the measures implemented during days 107-115 (April 16-25, 2002) can rapidly reduce the number of foreign input infected individuals to a very low level in one month.
- Different combinations of the eight parameters of Equation (1) may generate similar simulation results. Therefore need further study to obtain better parameter combinations to interpret COVID-19 epidemics.

A recommendation is that the administration should at least maintain the prevention and control measures implemented 7 days after reaching the turning point [15,16]. Figure 1 and Table 1 suggest that the importance of the recommendation.

It is not wise strategy to withdraw all prevention and control measures before the number of the all infected people have been cleared. 100% blocking the speed at which COVID-19 infection spreads is key Strategies for early clearance or reduction of epidemic spread possible [15, 16].

More strict prevention and control strategies are necessary to prevent the spread of COVID-19 with Omicron and Delta variations. It is expected that this research can provide better understanding, interpretation and leading the spread and control measures of epidemics.

## Data Availability

All data produced are available online at http://www.nhc.gov.cn/.

## Data availability statement

Data are available on reasonable request. Please email the author.

## Ethical Statement

Not applicable/No human participants included.

